# Sleep health interventions for managing mental health in shift workers: A systematic review

**DOI:** 10.1101/2025.07.28.25331984

**Authors:** Peter Bragge, Jane Burns, Paul Kellner, Monika Allan, David Fitzgerald, Emily Grundy, Alyse Lennox, Talar R. Moukhtarian, Shantha M.W. Rajaratnam, Tracey L. Sletten

## Abstract

Shift work can reduce sleep duration and quality, increasing the risk of mental health disorders which burden individuals, their families and productivity. This systematic review aimed to identify and characterise studies evaluating a sleep health intervention in shift workers and measuring a mental health outcome. Searching of seven academic databases identified 102 eligible studies. Interventions predominantly evaluated shift rostering (n=39); pharmacological interventions (n=25); and lighting (n=14). Almost half (n=43) of the included studies recruited healthcare workers. Effective sleep interventions for optimising mental health included forward-rotating (day, afternoon, night) shifts, restricting work hours, pharmaceutical agents and lighting interventions. Insights on the findings provided by two experienced shift workers identified relatively low awareness of the link between sleep and mental health, and variation in acceptability and feasibility of interventions between sectors. Business leaders and policymakers should work to increase awareness of sleep health in shift workers and consider tailored implementation of evidence-based interventions. Future research should prioritise under-represented populations, including those outside of the health and manufacturing sectors and in low- and middle-income countries, and consider expanding the evidence base beyond shift design, medications and lighting.

## 1. Introduction

While most people are sleeping, 15-20% of the working population in industrial societies are working across numerous sectors including manufacturing, hospitality services, emergency services and healthcare [1–3]. To provide these goods and services, shift workers often work variable patterns of early morning, afternoon/evening and overnight shifts. Shift work interferes with the human central circadian pacemaker [4] which promotes alertness during the day and sleepiness at night through the hormonal and nervous systems [5], for example via the nightly release of the hormone melatonin which facilitates sleepiness [4].

The human body cannot easily adapt the timing of the circadian pacemaker to the reversed pattern of work and sleep required by night shift [5]. The subsequent misalignment is associated with reduced quality and duration of sleep during the day and impaired alertness during night shift. These disruptions elevate risk of occupational and road accidents [5] and carry associated costs to society estimated at over US$50 billion/year for road accidents and US$20 billion/year for workplace accidents [6,7] Additionally, long-term disruptions to the circadian system have been linked to cardiovascular, metabolic, cognitive, reproductive and gastrointestinal disorders and certain cancers [3,6].

Circadian and sleep disturbances have also been associated with increased risk of negative mental health outcomes including depressive symptoms [1,8–10] and adverse impacts on social interaction, recreation and other ‘everyday’ activities. These impacts affect quality of life [1,6], further compromising mental health. Shift work therefore contributes disproportionately to the overall global burden of mental disorders, which make up over 14% of the global of burden of disease [1].

The adverse mental health impacts of working non-standard work hours highlight the need for interventions addressing sleep health in shift workers. These include pharmacological interventions [11], psychological therapies such as cognitive behavioural therapy (CBT) [12], and changes to the physical environment such as light therapy [13]. Research examining such interventions measures a range of outcomes including numerous measures of sleep quality and duration, productivity, cardiovascular and hormonal parameters. Although some studies also collect mental health data, mental health is often one of many outcomes examined in research examining sleep health interventions for shift workers. Therefore, the aim of systematic review was to comprehensively examine the effect of sleep health interventions for managing mental health in shift workers.

## 2. Methods

### 2.1 Protocol and registration

Systematic reviews are well-established for examining effectiveness and are the highest-ranked form of evidence for addressing intervention questions [14]. The review was developed in accordance with the PRISMA (Preferred Reporting Items for Systematic Reviews and Meta-Analyses) statement [15] and registered on the PROSPERO database in December 2021 (ref: CRD42022298879).

### 2.2 Search strategy

The search strategy was developed in collaboration with an information specialist at Monash University (Supplementary File 1). To refine the search strategy, we mapped known relevant studies citing studies using a web-based application (https://www.citationgecko.com/). We examined search yields from our test searches to ensure that they contained several studies from this ‘gold set.’ The Medline (OVID), Embase (OVID), Cochrane Central, CINAHL, PsycINFO and Business Source Complete, Institute for Work and Health (IWH) were then searched from inception to December 2021.

### 2.3 Inclusion and exclusion criteria

Due to uncertainty about the volume of evidence in this area, our review was inclusive in defining ‘sleep health interventions’ and also defined mental health broadly as *‘ … a state of well-being in which an individual realizes his or her own abilities, can cope with the normal stresses of life, can work productively and is able to make a contribution to his or her community’* [16]. This definition is not limited to absence of diagnosed mental illness.

Therefore, in addition to measures of depression, anxiety, stress and burnout, studies measuring mental health and mood outcomes were included in the review. This reflects the complex inter-relationships between shift work, sleep and mental health. Inclusion and exclusion criteria for the review are presented in Table 1.

**Table 1:** Inclusion and exclusion criteria for review.

|  | **Include** | **Exclude** |
| --- | --- | --- |
| **Study Type** | • Primary study examining effectiveness of sleep health interventions<br>• Trial protocols published since 2017<br>• Reviews published since 2017 (scanned to identify primary studies) | • Observational / qualitative / non-interventional studies<br>• Single case studies |
| **Population** | • Shift workers, defined as paid employees working non-standard hours which include evening, overnight or early morning including permanent shift work or rotating shift / standard work schedules<br>• Laboratory simulations of shift work (including in non-shift work participants) | • Non-working populations<br>• Non-shift workers (i.e., their main role is 9-5. They may do some overtime / out of hours but not routinely or mandated) |
| **Study Setting** | • No exclusions |  |
| **Study focus** | • Primary aim of study is establishing the effectiveness of a sleep health intervention in shift workers |  |
| **Outcome** | • All measures of mental health / mood | • No mental health / mood measures |
| **Publication Status** | • English language<br>• Peer-reviewed journal publications<br>• Reports from relevant Government / other institutions | • Dissertations<br>• Books / book chapters |

### 2.4 Study Selection

Search results were exported into Covidence (Cochrane technology platform). Title/abstracts and full text studies were independently screened by two reviewers drawn from the research team (PB, PK, TM, AL, EG, TS). Conflicts were resolved by a third reviewer (PB or PK).

### 2.5 Data extraction

Data extraction focused on the following data items: intervention type/subtype; study type; population (workplace industry where relevant); sample size; intervention/comparator; mental health outcomes; and direction of evidence. Eligible studies were dichotomised by intervention into those using a recognised mental health outcome (defined as those which referenced the mental health tool(s) used) and those which did not. The presence of a statistically significant finding pertaining to any sleep parameter was also recorded.

### 2.6 Involvement of experienced shift workers

The review team included two experienced shift workers recruited through a sleep researcher (TS) using a position description (Supplementary File 2) - a registered nurse with 14 years’ experience including several years working night shifts in a major metropolitan hospital (MA); and a health and safety manager with 25 years’ experience of shift work in the mining and construction sector (DF). The experienced shift workers participated in development of the review protocol (MA); provided reflections on the interventions identified in the review; and described strategies they used for managing their sleep.

## 3. Results

### Study selection

Searching identified 7,006 citations; 4,900 citations and abstracts were screened; 443 studies were assessed in full text; and 102 primary studies met the inclusion criteria (Supplementary File 3). Interventions identified were rostering (n=39); pharmacotherapies (n=25); lighting (n=13); cognitive behavioural therapy (CBT; n= 5); napping (n=5); and education/training (n=3). Eleven studies were either combinations of these or were single studies of an intervention. Healthcare workers were the most studied group (n=44). Fourteen studies were laboratory-based shift work simulations. Other participant groups included manufacturing workers (n=11); emergency workers (n=9); aviation workers (n=5); military personnel (n=3); and workers in extractive industries (n=3). Few studies focused only on workers identified as having insomnia (n=4) or shift work disorder (n=3). Tables 2 - 8 present key study characteristics and findings which are summarised below including experienced shift worker perspectives.

### Rostering interventions (N=39)

Most studies (32/39) used a referenced measure of mental health. Interventions encompassed changes to shift duration; shift cycle/rotation direction (forwards vs. backwards rotation) and on-call arrangements (Table 2). A *forward shift system* refers to shifts that rotate to a later time through a working week (e.g., from a morning to evening to night shift). Study designs were mainly pre-post (n=18) and quasi-experimental (n=9) rather than randomised.

**Table 2:** Summary of studies implementing rostering interventions (n=39)

| **First Author, Year** | **Study type** | **Population Category** | **Total n participants**<br>Group(s) (description of intervention): n | **Mental health outcomes (Sig in bold)** | **Sig. mental health outcome** | **Sig. sleep outcome** |
| --- | --- | --- | --- | --- | --- | --- |
| **Studies using a referenced mental health outcome measure (n = 32)** |  |  |  |  |  |  |
| Amendola et al. 2011 [21] | RCT | Police / Fire / other social services | **231**<br>C (8-h shift): 69<br>I1 (10-h shift): 81<br>I2 (12-h shift): 81 | **Quality of Life** | I1 Y+<br>I2 N | I1 Y+<br>I2 N |
| Antiel et al. 2013 [28] | Longitudinal | Healthcare workers | **156**<br>Duty hour regulations | Quality of Life;<br>Wellbeing;<br>Burnout; Job satisfaction | N | N |
| Bell et al. 2015 [27] | Quasi-experimental | Police / Fire / other social services | **386**<br>C (4x10-h shifts/wk): 197<br>I (3x13.3-h shifts/wk): 189 | **Stress; Quality of Life** | Y- | Y- |
| Bilimoria et al. 2016 [29] | Cluster randomised trial | Healthcare workers | **151 hospitals**<br>C (standard duty hours): 71<br>I (flexible duty hours): 80 | Wellbeing | N | N/A |
| Brainch et al. 2018 [30] | Pre-post | Healthcare workers | **10**<br>Change from 12-h to 10-h shifts | **Stress** | Y+ | Y+ |
| Caputo et al. 2015 [31] | Pre-post | Police / Fire / other social services | **59**<br>48-h on, 96-h off | Burnout;<br>Quality of Life | N | Y+ |

| First Author, Year | Study type | Population Category | Total n participants<br>Group(s) (description of intervention): n | Mental health outcomes<br>(Sig in bold) | Sig. mental health outcome | Sig. sleep outcome |
| --- | --- | --- | --- | --- | --- | --- |
| Chang et al. 2011 [32] | RCT | Healthcare workers | **62**<br>C (2 consecutive night shifts): 21<br>I1 (3 consecutive night shifts): 20<br>I2 (4 consecutive night shifts): 21 | Anxiety | I1 N<br>I2 N | I1 N<br>I2 N |
| Chang et al. 2018 [33] | Quasi-experimental case control study | Healthcare workers | **42**<br>C (off duty): 21<br>I: (two consecutive night shifts followed by 24-h off): 21 | Anxiety | N | Y+ |
| Hakola et al. 2021 [34] | Pre-post | Aviation workers; aircraft inspectors | **23**<br>I1 (12 mo. after change from forward rotating to fast forward rotating shift): 10<br>I2 (5 mo. after change from forward rotating to fast forward rotating shift): 13 | **Stress** | I1 Y+<br>I2 Y+ | I1 Y+<br>I2 Y+ |
| Hanoa et al. 2011 [35] | Pre-post | Extractive industries | **407**<br>C (7d on/7d off schedule in 2006): 226<br>I1 (7/7 in 2006, 7/7 in 2008): 44<br>I2 (7/7 in 2006, 14/14 in 2008): 137 | Stress;<br>**Coping** | I1 Y-<br>I2 Y+ | I1 N<br>I2 N |
| Harma et al. 2006 [22] | Quasi-experimental | Other shift workers; Maintenance | **140**<br>C (standard shift system): 116<br>I (rapid forward rotating shift system): 24 | **Wellbeing** | Y+ | Y+ |
| Kandolin & Huida 1996 [36] | Quasi-experimental | Healthcare workers | **58**<br>C (standard shift system): 13<br>I (clockwise shift rotating and planning shift 6 mo. in advance): 45 | **Mental strain; Stress** | Y+ | Y+ |
| Karlson et al. 2009 [37] | Before-after with daytime worker control group | Manufacturing | **185**<br>C (standard daytime workers): 67<br>I (change from forward- to backward-rotating schedule): 118 | **Stress;**<br>**Quality of Life** | Y+ | Y+ |
| Klein Hesselink et al. 2010 [18] | Before-after with control group | Manufacturing | **1583**<br>C: 399<br>I: (backward to forward + slow to fast rotating + extra day off after night shift): 1184 | **Stress** | Y+ | N/A |
| Knauth & Hornberger 1998 [38] | Controlled intervention study. 2 experimental and 2 control groups | Manufacturing | **179**<br>C1 (standard *discontinuous* shift): 34<br>I1 (from *discontinuous* weekly backward to a fast-forwarding *discontinuous* shift): 40<br>C2 (standard *continuous* shift): 54<br>I2 (from weekly backward to fast-forwarding *continuous* shift): 51 | **Quality of Life** | I1: Y+<br>I2: Y+ | I1: N<br>I2: N |
| Landrigan et al. 2008 [39] | Pre-post | Healthcare workers | **220**<br>Work-hour limits (ACGME standards): 220 | Depression;<br>Burnout | N | N |
| Loudoun 2008 [40] | Quasi-experimental | Manufacturing | **137**<br>C (standard 12-h shift): 17<br>I (shift increased from 8 to 12 h): 120 | Quality of Life | N | N |
| Low et al. 2018 [41] | Quasi-experimental (2 timepoints) | Healthcare workers | **43 timepoint 1; 27 timepoint 2**<br>C (traditional on-call: 4-6 night shifts monthly): 14 timepoint 1; 10 timepoint 2<br>I (night float: 5 consecutive nights per week bimonthly): 29 timepoint 1; 17 timepoint 2 | Quality of Life | N | N |
| Lowden et al. 1998 [42] | Pre-post | Manufacturing | **46**<br>C (8-h shifts): 14<br>I (two 12-h shifts): 32 | **Quality of Life; Psychological fatigue** | Y+ | Y+ |
| Mitchell & Williamson 2000 [43] | Pre-post | Extractive industries | **27**<br>Pre (8-h shifts): 15<br>Post (12-h shifts): 12 | **Mood; Stress** | Y+ | Y+ |
| Monk et al. 2006 [44] | Pre-post | Healthy volunteers | **10**<br>I: 12 x 30-minute phase advances in sleep schedule | **Mood; Alertness** | N | Y- |
| Monk et al. 2004 [45] | Pre-post | Healthy volunteers | **10**<br>I: 9 x 2h delays in sleep schedule | **Mood; Alertness** | N | Y- |
| Paley et al. 1998 [46] | Within-subjects repeated measure | Police/Fire/other social services | **24 (overlap between the two groups)**<br>Initial schedule (IS): 8-h night / afternoon / morning rotating backwards x 2 weeks: 20<br>**New schedule (NS): 10-h day / 14-h night x forwards x 2 days then 4 days off: 19**<br>15 provided data on both IS and NS | Mood | NS (night vs. day): Y-<br>NS vs. IS: N | NS (night vs. day): Y-<br>NS vs. IS: N |
| Parshuram et al. 2015 [47] | Pre-post | Healthcare workers | **47**<br>Pre: standard shift<br>Post1 (12-h shift): 17<br>Post2 (16-h shift): 15<br>Post3 (24-h shift): 12 | Burnout | N | N |
| Parthasarathy et al. 2007 [48] | Pre-post | Healthcare workers | **44**<br>Pre: standard shift (can be > 30 h/week)<br>Post: shift hour reduced to < 30 h/week | **Quality of Life** | Y+ | Y+ |
| Pierce & Dunham 1992 [49] | Pre-post | Police/Fire/other social services | **74**<br>Pre: fast forwarding shift (8-10-h shift)<br>Post: fast forwarding shift (12-48-h shift) | **Stress** | Y+ | Y+ |
| Ripp et al. 2015 [50] | Pre-post | Healthcare workers | **239**<br>Pre: 24-h shift<br>Post: 14 or 16 h shift | Burnout | N | N |
| Schuh et al. 2011 [23] | Pre-post | Healthcare workers | **23 (burnout); 29 (sleep)**<br>Pre & post IOM recommendations | **Burnout** | Y- (1 / 3 sub scores) | N |
| Shochat et al. 2019 [51] | Pre-post | Aviation workers | **39**<br>Pre: 8-h shift<br>Post: 12-h shift | **Burnout** | Y+ | Y+ |
| Sussman & Paul 2015 [52] | Pre-post | Healthcare workers | **56**<br>Pre: 24-h shift<br>Post: 16-h shift | Quality of Life; Burnout | N<br>(descriptive stats improved) | N/A |
| Vetter et al. 2015 [53] | Pre-post | Manufacturing | **114**<br>Pre: standard shift<br>Post: a chronotype-adjusted shift | **Wellbeing;**<br>Stress | Y+ | Y+ |
| Yamada et al. 2001 [54] | Pre-post with control | Manufacturing | **205**<br>Pre (8-h shift): 16<br>Post (moved from 8-h to 12-h shift): 189 | Mood | Y- | N/A |
| **Studies not using a referenced mental health outcome measure (n = 7)** |  |  |  |  |  |  |
| Auger et al. 2012 [25] | Partially randomised cohort study | Healthcare workers | **27** (3 in both C & E)<br>C (30-h shift every fourth night): 16<br>E (max 12-h shift): 14 | Well-being | N | Y |
| Cull et al. 2006 [17] | Repeated cross-sectional surveys pre and post policy implementation | Healthcare workers | **626 Residents**<br>Time 1 (before work hour limit): 323<br>Time 2 (after work hour limit): 303<br>+ 161 Program Directors (not statistically analysed) | **Mood** | Y+ | Y+ |
| De Valck et al. 2007 [55] | Quasi-experimental | Manufacturing | **38**<br>C (slow backward rotating shift): 18<br>E (fast forward rotating shift): 18 | Stress | N | Y+ |
| Duplessis et al. 2007 [56] | Single-factor (Schedule), 3-level design with repeated measures (essentially a 3-armed pre-post) | Military | **40**<br>Pre: backward rotating 6-h shift<br>Post: fast-forward rotating 6-h shift | Stress | N | Y+ |
| Fitzgibbons et al. 2012 [24] | Yearly cross-sectional surveys | Healthcare workers | **216**<br>Survey: 8 years after participating in an hour limit program | **Satisfaction** | Y- | N |
| Kamine et al. 2013 [57] | Pre-Post | Healthcare workers | **30**<br>**ACGME implementation**<br>Pre: overnight shift every 4<sup>th</sup> night<br>Post: a night float schedule | **Mood (days cried);** Mood (all other) | Y- | N/A |
| Miulli & Valcore 2010 [20] | Post intervention survey | Healthcare workers | **108**<br>post-intervention survey | Quality of Life | N | N |

ACGME = Accreditation Council for Graduate Medical Education; C = control group; d = Days; I = intervention group; IOM = Institute of Medicine; Mo. = Months; n = number of participants; N = No statistically significant result (i.e. no difference between groups detected); N/A = Not applicable (i.e. not measured); RCT = randomised controlled trial; Sig. = Statistically significant; Y = Statistically significant result, where + favours intervention and – favours control

Populations were predominantly from healthcare (n=19); manufacturing (n=8); police / fire and social services (n=5) and extractive industries. In contrast to other interventions, almost half of the studies (n=17) had over 100 participants. Mental health outcomes were evenly spread with at least 5 studies examining burnout, wellbeing, stress and quality of life. Twelve studies reported positive effects including six with samples of over 100. All except one [17] used referenced mental health outcome measures.

In 1583 manufacturing workers, positive impacts on work stress were reported in response to a change from backward to forward rotating rosters; reduction from three to two ‘fast rotating’ shifts; and three instead of two days off after night shifts [18]. Switching from a backward to a forward shift system was also associated with improvements in quality of life amongst 179 steel manufacturing workers [19].

- Cull et al. (2006) examined the impact of new Graduate Medical Education resident duty hour limits, reporting improved resident self-reported well-being in a sample of 767 program directors and graduating residents. [17] Similarly, Miulli et al.’s 2010 study of 108 residents reported positive impacts; [20]
- Amendola et al. (2011) reported a significant effect of shift length on quality of work life in 231 police, with the highest quality of work life when working 10-hour shifts; [21]
- Knauth et al. (1998) reported improvements in quality of life amongst 179 steel manufacturing workers following a switch from a backward to a forward shift system; [19] and
- Harma et al. (2005) examined rapid forward-rotating shifts which avoided consecutive night shifts and optimised free time in 140 airline workers, reporting positive well-being outcomes. [22]

Another nine studies reflected the above findings on the impact of duty hour limits; however, Schuh et al. (2011) [23] have reported a negative effect of the Institute of Medicine work hours restrictions on burnout in 34 medical residents.

Twelve studies reported no effect of shift manipulations on mental health outcomes. Five studies reported negative outcomes; however these were not restricted to wellbeing. One study by Fitzgibbons et al. (2012) [24] reported from 216 survey responses that duty hour restrictions had led to a non-significant improvement in wellbeing but also a significant decrease in satisfaction with medical education. Increased work compression and fatigue resulting from duty hour restrictions in 27 residents was reported by Auger et al. (2012) [25], however, neither of these studies used a referenced mental health outcome. Another two studies reported negative effects including weight gain and fatigue following a change from 8-hour to 12-hour shifts in 205 cleanroom workers [26] and negative effects of long (over 13- hour) shifts on quality of life in 386 police officers [27].

Collectively, these findings indicate that shifting to a forward rotating shift schedule has a generally positive impact on mental health across multiple professions; work hour restrictions in resident-level doctors have positive mental health impacts but a perception of compromised professional education; and shorter (10-hour) shifts are optimal compared with longer (12+ hour) shifts. The existence of studies with generally higher sample sizes conducted across multiple disciplines indicates that comparatively more confidence can be placed in these findings compared to other intervention categories.

### Experienced shift worker reflections: rostering

In mining and construction, the mistaken belief that people ‘adapt’ their circadian system to night shifts leads some workers to prefer many consecutive night shifts even though this adversely affects health, and increased pay for night shift may also drive this preference.

Combined with the fact that some construction tasks cannot be done at night, this raises the question of whether night shifts in construction represent good return on investment.

Conversely, there is no question of the need for shift work in nursing. Systems for rostering include online tools that enable nurses to indicate availability over a defined period. This assists managers in the considerable task of coordinating multiple staff and having some control over rostering may contribute to lower work-related stress in employees since higher occupational autonomy is associated with better wellbeing and workplace productivity. Ten-hour shifts appear more frequently in nursing compared to mining and construction, where 12-hour shifts can occur, especially in large construction projects. Across both these sectors (and potentially others), a lack of research-informed education may contribute to low employee awareness of the potential impacts of shift work on mental health and work-life balance.

### Pharmacological interventions (n=25)

Most studies examining pharmacological interventions (23/25) used a referenced measure of mental health and employed a randomised controlled trial or crossover-RCT design (Table 3). The most frequently studied agents were melatonin and other hypnotics to target improvements in sleep; and stimulants (e.g., modafinil, armodafinil) to target sleepiness when awake. Only two studies recruited over 100 participants. Fourteen of the pharmacological studies reported positive effects on mood.

**Table 3:**
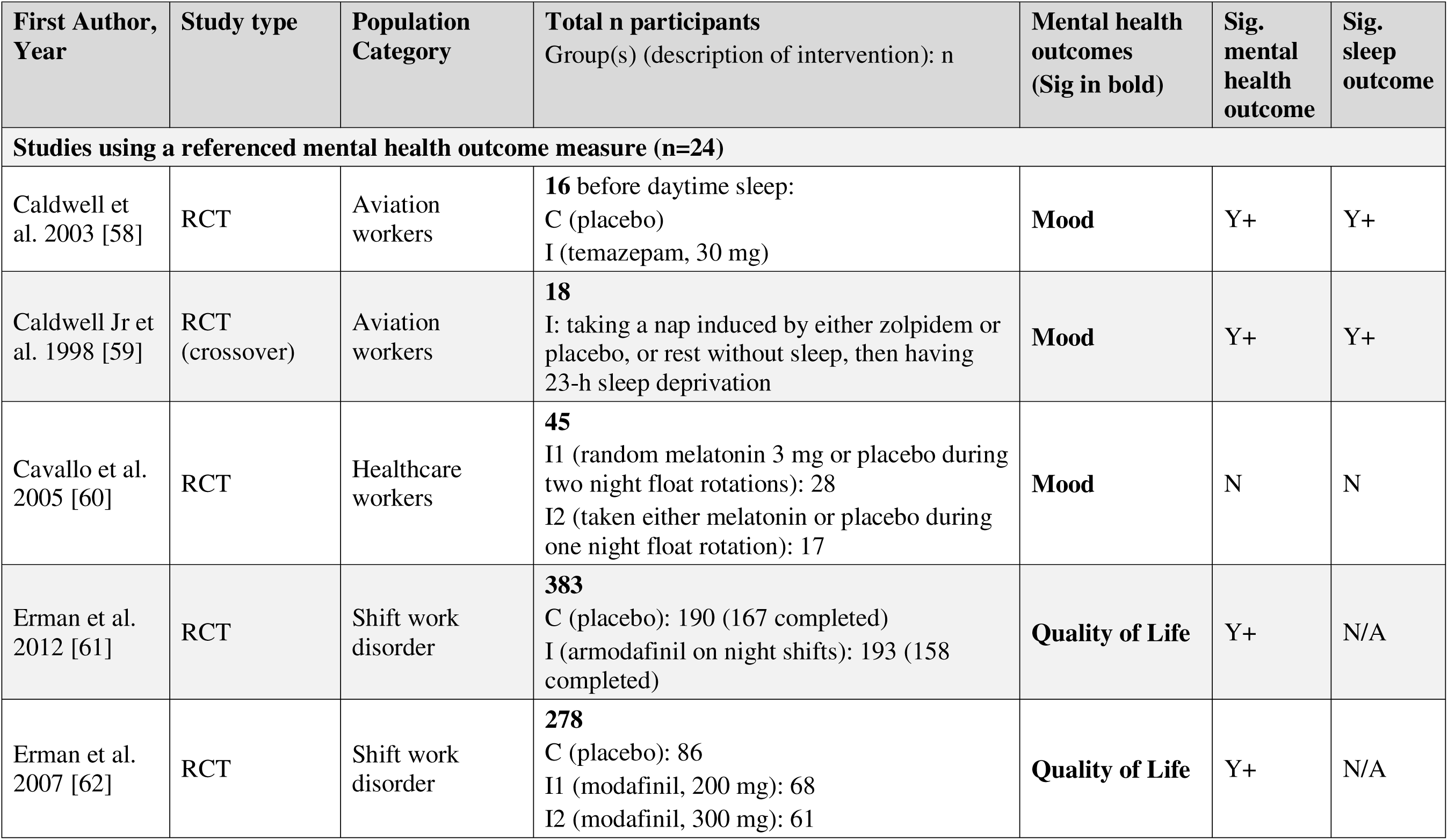
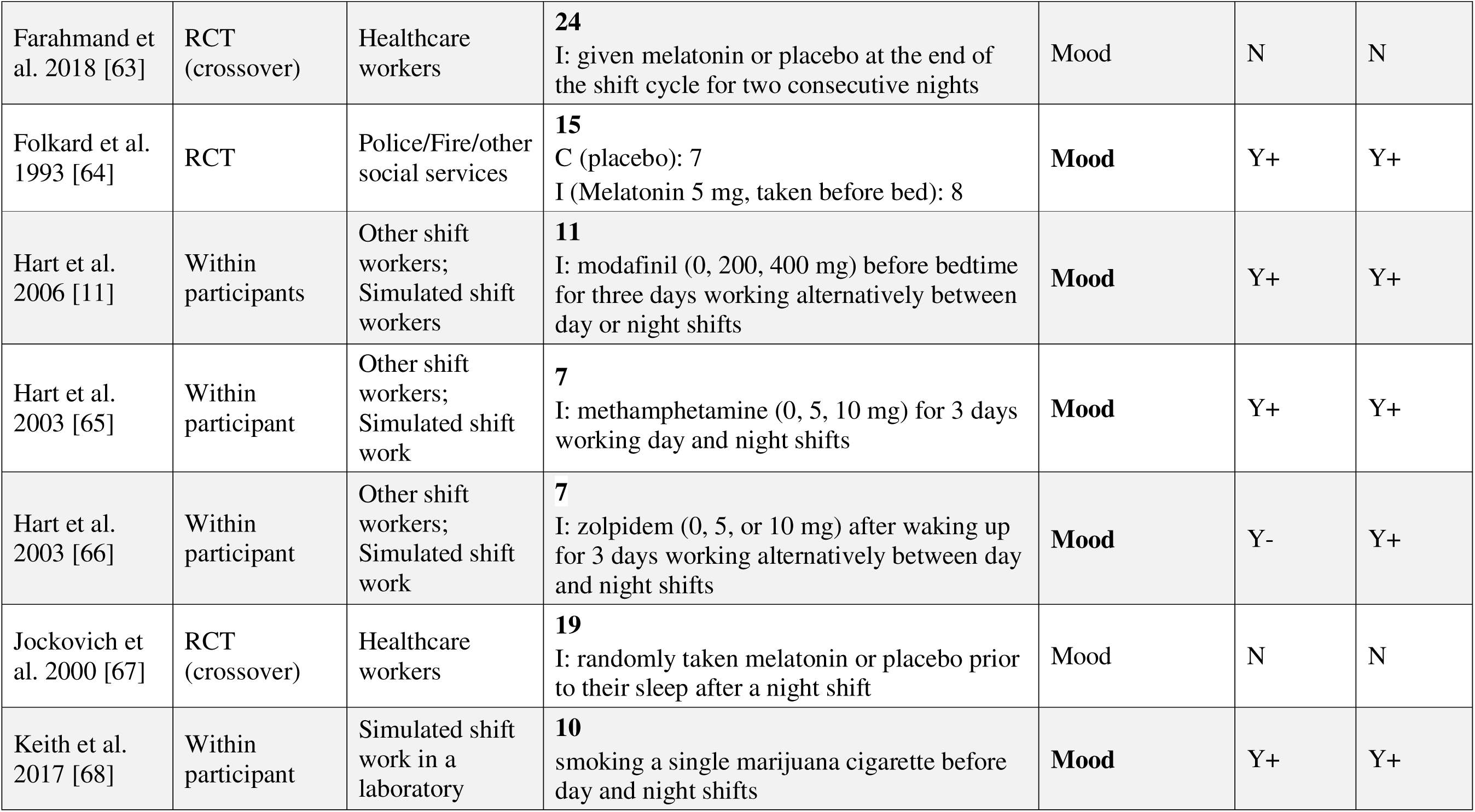
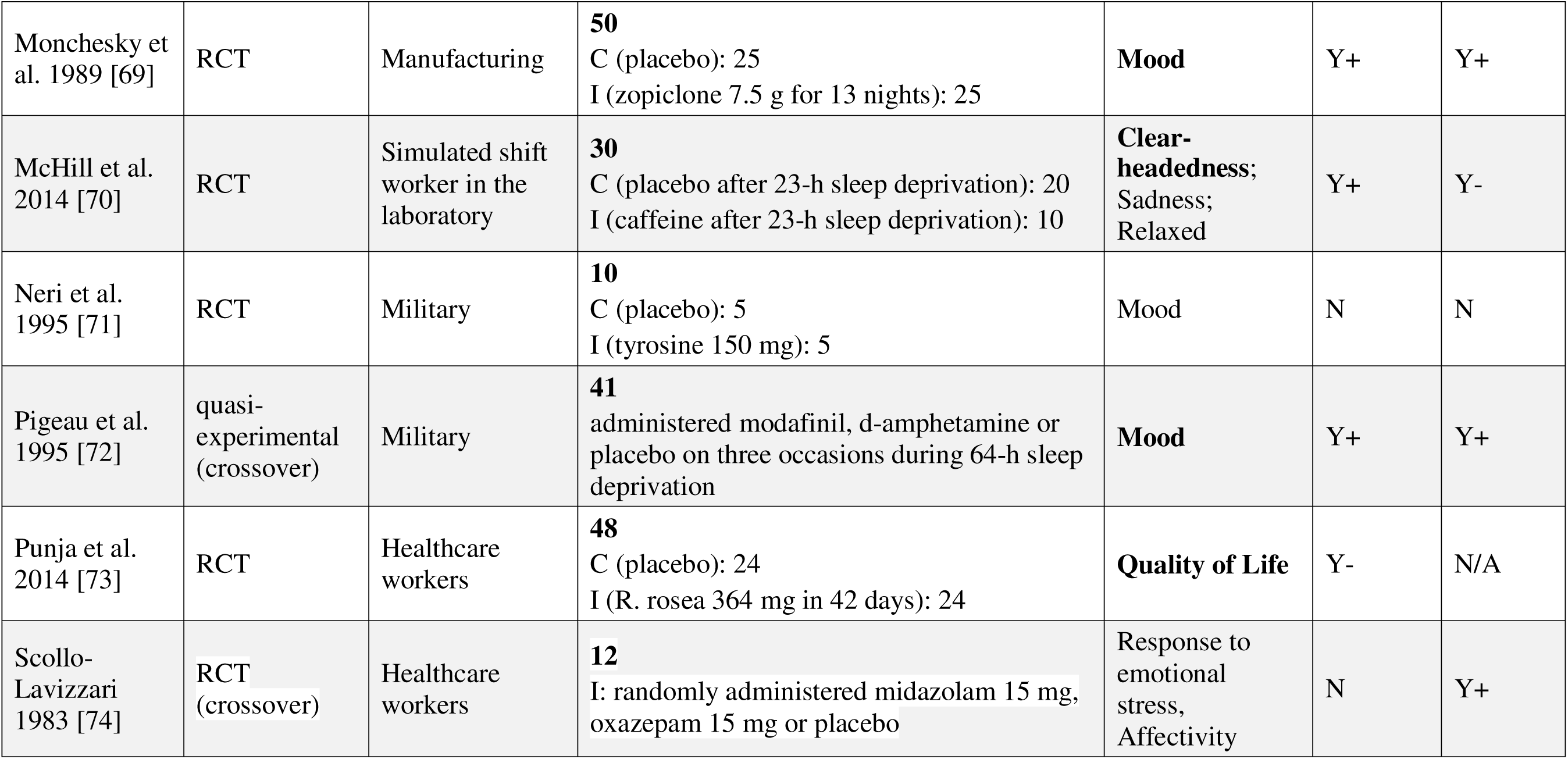
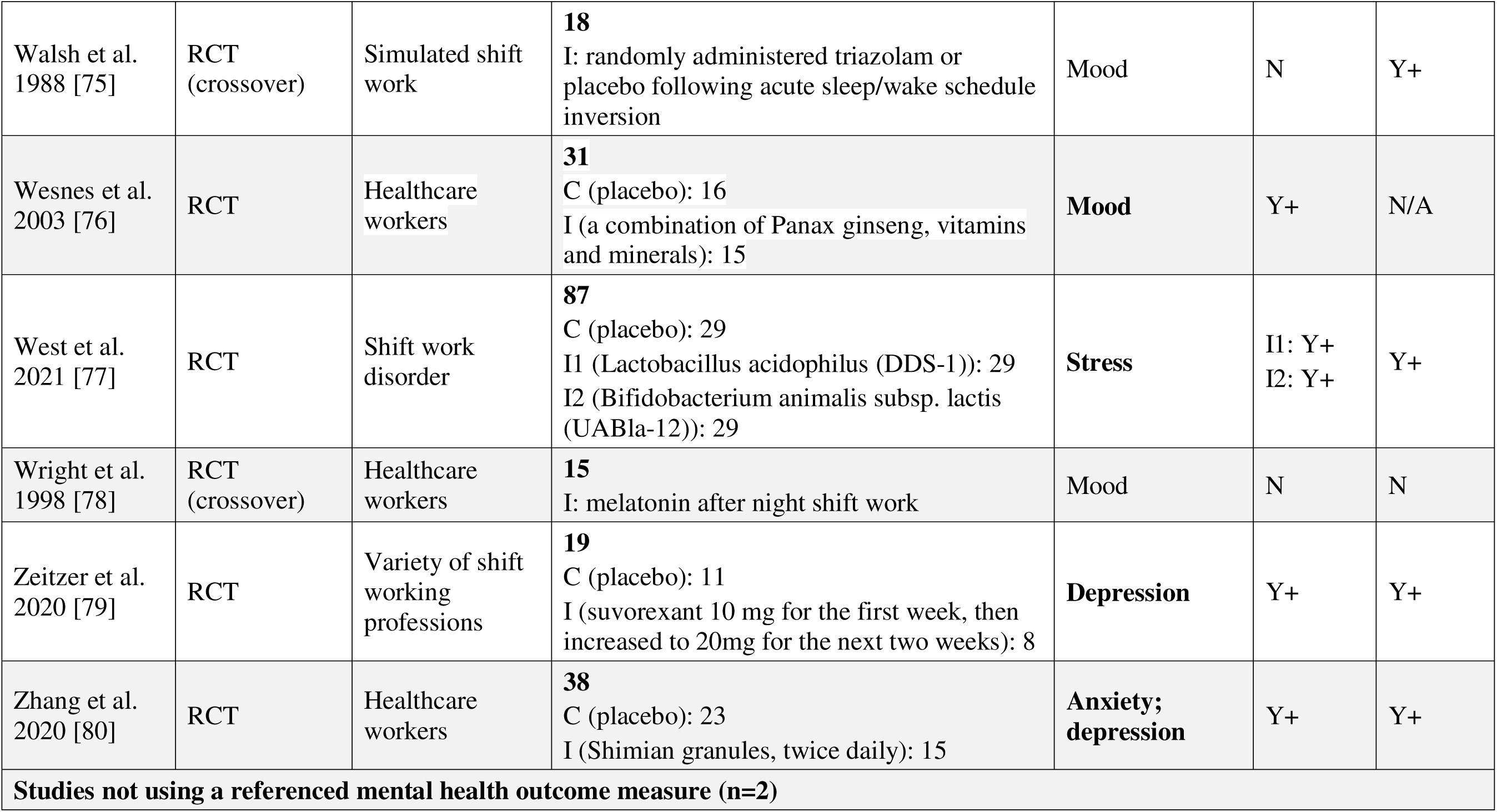
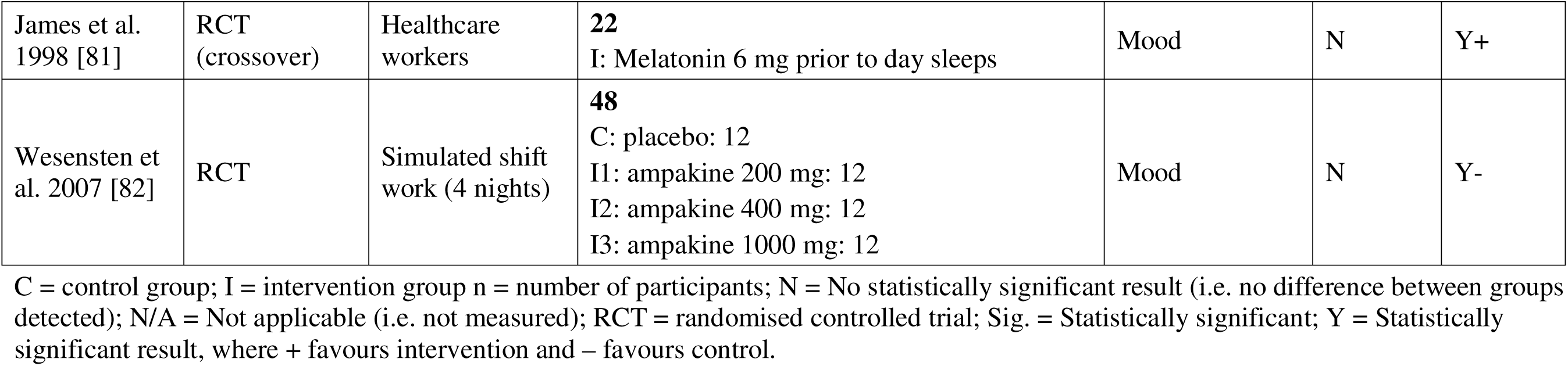
Summary of studies implementing pharmacological interventions (n = 26)

| **First Author, Year** | **Study type** | **Population Category** | **Total n participants**<br>Group(s) (description of intervention): n | **Mental health outcomes**<br>(Sig in bold) | **Sig. mental health outcome** | **Sig. sleep outcome** |
| --- | --- | --- | --- | --- | --- | --- |
| **Studies using a referenced mental health outcome measure (n=24)** |  |  |  |  |  |  |
| Caldwell et al. 2003 [58] | RCT | Aviation workers | **16** before daytime sleep:<br>C (placebo)<br>I (temazepam, 30 mg) | **Mood** | Y+ | Y+ |
| Caldwell Jr et al. 1998 [59] | RCT (crossover) | Aviation workers | **18**<br>I: taking a nap induced by either zolpidem or placebo, or rest without sleep, then having 23-h sleep deprivation | **Mood** | Y+ | Y+ |
| Cavallo et al. 2005 [60] | RCT | Healthcare workers | **45**<br>I1 (random melatonin 3 mg or placebo during two night float rotations): 28<br>I2 (taken either melatonin or placebo during one night float rotation): 17 | **Mood** | N | N |
| Erman et al. 2012 [61] | RCT | Shift work disorder | **383**<br>C (placebo): 190 (167 completed)<br>I (armodafinil on night shifts): 193 (158 completed) | **Quality of Life** | Y+ | N/A |
| Erman et al. 2007 [62] | RCT | Shift work disorder | **278**<br>C (placebo): 86<br>I1 (modafinil, 200 mg): 68<br>I2 (modafinil, 300 mg): 61 | **Quality of Life** | Y+ | N/A |

| First Author, Year | Study type | Population Category | Total n participants<br>Group(s) (description of intervention): n | Mental health outcomes<br>(Sig in bold) | Sig. mental health outcome | Sig. sleep outcome |
| --- | --- | --- | --- | --- | --- | --- |
| Farahmand et al. 2018 [63] | RCT (crossover) | Healthcare workers | **24**<br>I: given melatonin or placebo at the end of the shift cycle for two consecutive nights | Mood | N | N |
| Folkard et al. 1993 [64] | RCT | Police/Fire/other social services | **15**<br>C (placebo): 7<br>I (Melatonin 5 mg, taken before bed): 8 | **Mood** | Y+ | Y+ |
| Hart et al. 2006 [11] | Within participants | Other shift workers;<br>Simulated shift workers | **11**<br>I: modafinil (0, 200, 400 mg) before bedtime for three days working alternatively between day or night shifts | **Mood** | Y+ | Y+ |
| Hart et al. 2003 [65] | Within participant | Other shift workers;<br>Simulated shift work | **7**<br>I: methamphetamine (0, 5, 10 mg) for 3 days working day and night shifts | **Mood** | Y+ | Y+ |
| Hart et al. 2003 [66] | Within participant | Other shift workers;<br>Simulated shift work | **7**<br>I: zolpidem (0, 5, or 10 mg) after waking up for 3 days working alternatively between day and night shifts | **Mood** | Y- | Y+ |
| Jockovich et al. 2000 [67] | RCT (crossover) | Healthcare workers | **19**<br>I: randomly taken melatonin or placebo prior to their sleep after a night shift | Mood | N | N |
| Keith et al. 2017 [68] | Within participant | Simulated shift work in a laboratory | **10**<br>smoking a single marijuana cigarette before day and night shifts | **Mood** | Y+ | Y+ |

| **First Author, Year** | **Study type** | **Population Category** | **Total n participants**<br>Group(s) (description of intervention): n | **Mental health outcomes (Sig in bold)** | **Sig. mental health outcome** | **Sig. sleep outcome** |
| --- | --- | --- | --- | --- | --- | --- |
| Monchesky et al. 1989 [69] | RCT | Manufacturing | **50**<br>C (placebo): 25<br>I (zopiclone 7.5 g for 13 nights): 25 | **Mood** | Y+ | Y+ |
| McHill et al. 2014 [70] | RCT | Simulated shift worker in the laboratory | **30**<br>C (placebo after 23-h sleep deprivation): 20<br>I (caffeine after 23-h sleep deprivation): 10 | **Clear-headedness;**<br>Sadness;<br>Relaxed | Y+ | Y- |
| Neri et al. 1995 [71] | RCT | Military | **10**<br>C (placebo): 5<br>I (tyrosine 150 mg): 5 | Mood | N | N |
| Pigeau et al. 1995 [72] | quasi-experimental (crossover) | Military | **41**<br>administered modafinil, d-amphetamine or placebo on three occasions during 64-h sleep deprivation | **Mood** | Y+ | Y+ |
| Punja et al. 2014 [73] | RCT | Healthcare workers | **48**<br>C (placebo): 24<br>I (R. rosea 364 mg in 42 days): 24 | **Quality of Life** | Y- | N/A |
| Scollo-Lavizzari 1983 [74] | RCT (crossover) | Healthcare workers | **12**<br>I: randomly administered midazolam 15 mg, oxazepam 15 mg or placebo | Response to emotional stress, Affectivity | N | Y+ |

| First Author, Year | Study type | Population Category | Total n participants<br>Group(s) (description of intervention): n | Mental health outcomes<br>(Sig in bold) | Sig. mental health outcome | Sig. sleep outcome |
| --- | --- | --- | --- | --- | --- | --- |
| Walsh et al. 1988 [75] | RCT (crossover) | Simulated shift work | **18**<br>I: randomly administered triazolam or placebo following acute sleep/wake schedule inversion | Mood | N | Y+ |
| Wesnes et al. 2003 [76] | RCT | Healthcare workers | **31**<br>C (placebo): 16<br>I (a combination of Panax ginseng, vitamins and minerals): 15 | **Mood** | Y+ | N/A |
| West et al. 2021 [77] | RCT | Shift work disorder | **87**<br>C (placebo): 29<br>I1 (Lactobacillus acidophilus (DDS-1)): 29<br>I2 (Bifidobacterium animalis subsp. lactis (UABla-12)): 29 | **Stress** | I1: Y+<br>I2: Y+ | Y+ |
| Wright et al. 1998 [78] | RCT (crossover) | Healthcare workers | **15**<br>I: melatonin after night shift work | Mood | N | N |
| Zeitzer et al. 2020 [79] | RCT | Variety of shift working professions | **19**<br>C (placebo): 11<br>I (suvorexant 10 mg for the first week, then increased to 20mg for the next two weeks): 8 | **Depression** | Y+ | Y+ |
| Zhang et al. 2020 [80] | RCT | Healthcare workers | **38**<br>C (placebo): 23<br>I (Shimian granules, twice daily): 15 | **Anxiety; depression** | Y+ | Y+ |
| **Studies not using a referenced mental health outcome measure (n=2)** |  |  |  |  |  |  |
| James et al. 1998 [81] | RCT (crossover) | Healthcare workers | **22**<br>I: Melatonin 6 mg prior to day sleeps | Mood | N | Y+ |
| Wesensten et al. 2007 [82] | RCT | Simulated shift work (4 nights) | **48**<br>C: placebo: 12<br>I1: ampakine 200 mg: 12<br>I2: ampakine 400 mg: 12<br>I3: ampakine 1000 mg: 12 | Mood | N | Y- |

C = control group; I = intervention group n = number of participants; N = No statistically significant result (i.e. no difference between groups detected); N/A = Not applicable (i.e. not measured); RCT = randomised controlled trial; Sig. = Statistically significant; Y = Statistically significant result, where + favours intervention and – favours control.

Several studies of hypnotics reported positive results for mood and mental health symptoms [58, 74, 79, 64]. Modafinil during shifts had significant positive effects on quality of life in an RCT of 278 participants [62]. It was also found to be a good alternative to amphetamine in a quasi-experimental study of 41 military personnel [72], and reversed disruptions in cognitive performance and mood during the night shift in a small, simulated shift work study [11]. Armodafinil had possible positive effects on quality of life after 6 weeks following an RCT in 383 participants [61]. Caffeine had positive effects on mood in a laboratory-based study of 30 participants [70]. Shimian granules (SMG, a herbal remedy) taken twice daily for a month improved anxiety and depression symptoms in 53 shift nurses [80]. Small studies (n under 20) reported positive effects of Marijuana [68], Methamphetamine [65], Temazepam [58] and Zolpidem [66].

These studies indicate that pharmacological agents can have positive effects on mental health in shift workers. Specifically, four studies reported that hypnotics positively impacted mood and mental health symptoms, and a further three studies, including the study with the largest sample size, reported positive effects of Modafinil. The remaining seven studies reporting positive impacts of pharmacological interventions all examined different agents, and therefore little confidence can be placed in these findings. Some adverse effects on mental health were reported for Zolpidem [66]; however, these were rare in the context of the overall body of evidence.

### Experienced shift worker reflections: pharmacological interventions

In mining and construction, pharmacological use is quite common, especially stimulants such as caffeine. Sedative agents are less frequently used, however there is a misconception that alcohol aids sleep. Understanding of the specific role of melatonin is relatively poor in these sectors, with many believing that it is a ‘sleeping tablet’. Another factor worthy of consideration in construction, being male-dominated, is the stigma associated with help-seeking for mental illness which may reinforce self-medication. Unlike in construction, melatonin and sedative use (e.g. Stilnox and Temazepam) may be more frequent in nursing. Echoing the above, education on appropriate use of medications and the potential harms associated with misuse could reduce adverse consequences associated with ‘self-medication’ with both pharmacological agents and other substances such as alcohol and caffeine.

### Lighting interventions (N=14)

Twelve of the 14 studies examining lighting interventions in shift work used a referenced measure of mental health (Table 4). Studies explored various combinations of duration of exposure and intensity of light to target alertness, mood and circadian phase shifting. Most studies employed an RCT crossover (n=5) or RCT (n=4) design, with the remainder pre-post (n=3) and quasi-experimental (n=2). Six studies recruited healthcare workers, including one with a focus on healthcare workers with insomnia. All sample sizes were under 100. The predominant mental health outcome examined in light studies was mood (n=10). Four studies reported positive findings for the effect of light on mood. Positive effects of lighting on wellbeing were also reported in two studies and anxiety and depression were reported to improve in one study. In most studies, however, mood was not the primary outcome of interest, with light primarily applied to influence sleep health through its influence on the timing or phase of the circadian rhythm, or acute alerting effects.

**Table 4:** Summary of studies implementing rostering lighting interventions (n = 13)

| **First Author, Year** | **Study type** | **Population Category** | **Total n participants**<br>Group(s) (description of intervention): n | **Mental health outcomes**<br>(Sig in bold) | **Sig. mental health outcome** | **Sig. sleep outcome** |
| --- | --- | --- | --- | --- | --- | --- |
| **Studies using a referenced mental health outcome measure (n=11)** |  |  |  |  |  |  |
| Canazei et al. 2017 [88] | RCT (crossover) | Simulated night shift in laboratory | **31**<br>Low, 2166 K (23:00–04:30 h)<br>Moderate, 3366 K<br>High, 4667 K | Mood | N | N |
| Costa et al. 1995 [83] | RCT (crossover) | Healthcare workers | **15**<br>C: no light intervention (100 lux)<br>I: 4 x 20 min of bright light (2,350 lux) during night duty | **Mood;**<br>Plasma cortisol | Y+ | Y+ |
| Costa et al. 1997 [84] | RCT (crossover) | Healthcare workers | **5**<br>C: no light visor<br>I: Light visor 4 x 40 min during 2 night shifts | Mood;<br>Plasma cortisol | N | N/A |
| Eastman 1992 [85] | Pre-post | Simulated night shifts at home | **24**<br>I1 (6-h light at same clock time every night for 4 nights): 18<br>I2 (6-h light, 1 h later each night for 4 nights): 6 | Mood | N | N |
| Eastman et al. 1995 [86] | Quasi-experimental | Simulated night shifts at home | **46**<br>C (<500 lux): 13<br>I1 (5,000 lux for 6 h): 16<br>I2 (5,000 lux for 3 h): 17 | **Mood** | Y+ | Y+ |

| First Author, Year | Study type | Population Category | Total n participants<br>Group(s) (description of intervention): n | Mental health outcomes<br>(Sig in bold) | Sig. mental health outcome | Sig. sleep outcome |
| --- | --- | --- | --- | --- | --- | --- |
| Eastman et al. 1994 [87] | Quasi-experimental | Simulated night shifts at home | **50**<br>C1 (dim light, no goggles): 13<br>C2 (dim light, goggles): 13<br>I2 (bright light, no goggles): 11<br>I3 (bright light, goggles): 13 | Mood | N | N |
| Canazei et al. 2017 [88] | RCT (crossover) | Simulated night shift in laboratory | **31**<br>Low, 2166 K (23:00–04:30 h)<br>Moderate, 3366 K<br>High, 4667 K | Mood | N | N |
| Comtet et al. 2019 [89] | RCT (crossover) | Simulated night shift in laboratory | **18**<br>C (ambient dim-light at 8 lux)<br>I1 (10,000 lux light box)<br>I2 (2,000 lux LED blue-enriched glasses)<br>30-min light exposure at 0500 h after simulated shift | Mood | N | N/A |
| Harrison et al. 2020 [13] | Pre-Post | Healthcare workers | **19**<br>Multi-component intervention:<br>6500 K architectural lighting in nurses' station plus optional behavioral components (light box, blue blocker glasses, eye mask) | **Quality of Life** | Y+ | Y |
| Huang et al. 2013 [90] | RCT | Healthcare workers; Insomnia | **92**<br>C (no bright light, sunglasses after work): 46<br>I (7-10,000 lux for $\geq 30$ min during the first half of the evening/ night shift): 46<br>[At least 10 days in 2 weeks] | **Anxiety; Depression** | Y+ | Y+ |

| **First Author, Year** | **Study type** | **Population Category** | **Total n participants**<br>Group(s) (description of intervention): n | **Mental health outcomes (Sig in bold)** | **Sig. mental health outcome** | **Sig. sleep outcome** |
| --- | --- | --- | --- | --- | --- | --- |
| Rahman et al. 2013 [91] | RCT (crossover) | Healthcare workers | **9**<br>C (standard indoor light)<br>I (short-wavelength light during night shift) | **Mood** | Y+ | Y+ |
| Sletten et al. 2021 [92] | RCT (crossover) | Manufacturing | **28**<br>C (4000 K, 43 lux during night shifts)<br>I (17000 K, 106 lux during night shifts) | Mood | N | N/A |
| **Studies not using a referenced mental health outcome measure (n=2)** |  |  |  |  |  |  |
| Figueiro et al. 2001 [93] | Pre-post | Healthcare workers | **21**<br>Bright light in nursing lounge room (2300-4000 lux on average) | Wellbeing;<br>Subjective wakefulness, motivation and feeling | N | N/A |
| Stewart et al. 1995 [86] | RCT | NASA personnel | **18**<br>C: 10<br>I (-10,000 lux for phase delay): 8 | Wellbeing;<br>**Irritability, sadness, anxiety** | Y+ | Y+ |

C = control group; I = intervention group K = kelvin; n = number of participants; N = No statistically significant result (i.e. no difference between groups detected); N/A = Not applicable (i.e. not measured); RCT = randomised controlled trial; Sig. = Statistically significant; Y = Statistically significant result, where + favours intervention and – favours control.

### Experienced shift worker reflections: lighting

In both mining and construction and nursing, most are unaware of the potential for light to aid sleep health and comparatively more attention is paid to other strategies (e.g., shift design). In some nursing settings there is a ‘night duty’ light switch in which environmental lighting is considerably dimmed. However, this is primarily directed towards promoting sleep for patients, whereas lighting design in this sector should also accommodate for brighter (rather than dim) lighting for workers given the links with light exposure and mental health. Healthcare is a somewhat unique sector in relation to lighting due to the need to accommodate two groups differently; workers and patients.

### Cognitive Behavioural Therapy (CBT) (N=5)

All five studies used a referenced measure of mental health (Table 5). CBT approaches included tailored sessions for specific workforces; for example, Jang (2020) [94] examined therapy for insomnia and nightmares in firefighters. Study populations were mixed and included healthcare and aviation workers. All five studies found statistically significant effects; however, all had sample sizes under 80. Despite the generally positive results, the variation in the content of CBT, populations and small sample sizes, makes it difficult to draw firm conclusions on the effect of CBT on mental health outcomes in shift workers.

**Table 5:** Summary of studies implementing Cognitive Behavioural Therapy (CBT) (n= 5)

| **First Author, Year** | **Study type** | **Population Category** | **Total n participants**<br>Group(s) (description of intervention): n | **Mental health outcomes (Sig in bold)** | **Sig. mental health outcome** | **Sig. sleep outcome** |
| --- | --- | --- | --- | --- | --- | --- |
| **Studies using a referenced mental health outcome measure (n=5)** |  |  |  |  |  |  |
| Jang et al. 2020 [94] | Pre-post | Police/Fire/other social services | **39**<br>CBT for insomnia and Imagery Reversal Therapy | **Depression; PTSD** | Y+ | Y+ |
| Järnefelt et al. 2012 [12] | Pre-post | Other shift workers; Insomnia | **19**<br>CBT for insomnia | **Quality of Life; Depression; Anxiety** | Y+ | Y+ |
| Järnefelt et al. 2020 [95] | "Partially randomised" | Healthcare workers; Other shift workers; Aviation workers; Police/Fire/other social services; airline, bakery, media; Insomnia | **75**<br>C (Sleep hygiene education): 22<br>I1 (Group CBT): 26<br>I2 (Self-help CBT): 27 | Depression; Quality of Life; Burnout; Stress; **Worry; Global effectiveness in improving their overall life situations and well-being** | Y+ | N/A |
| Lee et al. 2014 [96] | Prospective longitudinal within-subjects | Healthcare workers | **21**<br>C (4-week active control)<br>I (4-week CBT for shift workers) | **Depression; Anxiety** | Y+ | Y+ |
| Peter et al. 2019 [97] | Non-randomised trial | Manufacturing; Shift work disorder | **33**<br>I1 (face-to-face CBT): 12<br>I2 (online CBT): 21 | **Depression; Wellbeing** | Y+ | Y + |

C = control group; CBT = cognitive behavioural therapy; I = intervention group; n = number of participants; N = No statistically significant result (i.e. no difference between groups detected); N/A = Not applicable (i.e. not measured); PTSD = posttraumatic stress disorder; RCT = randomised controlled trial Sig. = Statistically significant; Y = Statistically significant result, where + favours intervention and – favours control.

### Experienced shift worker reflections: CBT

In mining and construction, CBT specific to the issue of sleep health is not frequently encountered, with mental health being a much greater focus (with passing references to rest, sleep and mindfulness). Similarly, the framing of CBT in nursing should emphasise its application to sleep health, which is the focus of most of the included studies. Although this type of CBT does have ‘flow-on’ benefits to mental health, it is primarily directed at sleep health behaviours. This means that participants do not need to identify as suffering from mental health challenges to participate. This subtle distinction is important as negative connotations can be associated with therapies for managing mental health.

### Napping interventions (N=5)

All five napping studies used a referenced measure of mental health (Table 6). Four of the five studies were RCTs, with one crossover-RCT design. Two studies recruited healthcare workers; two were laboratory-based simulations and one study recruited air traffic controllers. Naps were between 10 and 120 minutes. Mental health outcomes measured were mood (n=3) and anxiety (n=2). Results of napping on mood were mixed; one study reported a negative effect of a 10-minute nap on mood compared with a longer 30-minute nap (n=31, laboratory-based) [98], one reported more vigour with a 40-minute nap compared with no-nap (n=49 healthcare workers) [101], and one study reported no effect on mood of either a 45-minute or 120-minute nap (n=51 aviation workers) [100]. Of the two studies measuring anxiety, one reported no effect of a 30-min nap (n=63 healthcare workers) [99] and the other found significant reversal of anxiety following a 120-minute nap (n=13, laboratory-based) [102]. Given the modest sample sizes and variation in participant groups and nap times, no firm conclusions on the effect of napping on mental health outcomes in shift workers can be made from the included studies.

**Table 6:** Summary of studies implementing napping interventions (n = 5)

| First Author, Year | Study type | Population Category | Total n participants<br>Group(s) (description of intervention): n | Mental health outcomes<br>(Sig in bold) | Sig. mental health outcome | Sig. sleep outcome |
| --- | --- | --- | --- | --- | --- | --- |
| **Studies using a referenced mental health outcome measure (n = 5)** |  |  |  |  |  |  |
| Centofanti et al. 2016 [98] | RCT | Simulated night shift in laboratory | **31**<br>C (no nap): 11<br>I1 (10-min nap): 10<br>I2 (30-min nap): 10 | **Mood** | I1 Y- | N/A |
| Chang et al. 2015 [99] | RCT | Healthcare workers | **63**<br>C1 (Day shift): 21<br>C2 (Night shift without a nap): 21<br>I (Night shift with a 30-min nap): 21 | Anxiety | N | N/A |
| Della Rocco et al. 2000 [100] | RCT | Aviation workers | **51**<br>C1 (no nap): 16<br>I1: (45-min nap): 18<br>I2 (2-h nap): 17 | Mood | N | N/A |
| Smith-Coggins et al. 2006 [101] | RCT | Healthcare workers | **49**<br>C (no nap): 23<br>I (40-min nap): 26 | **Mood: Vigor**, tension, depression, confusion, anger | Y+ | N/A |
| Takeyama et al. 2002 [102] | RCT (crossover) | Simulated shift work | **13** [Mod. morning-types: 5, Mod. evening-types: 8]<br>C (no nap)<br>I (2-h nap) | Salivary cortisol (stress);<br>**Anxiety** | Y+ (morning types) | N/A |

C = control group; I = intervention group; n = number of participants; N = No statistically significant result (i.e. no difference between groups detected); N/A = Not applicable (i.e. not measured); RCT = randomised controlled trial Sig. = Statistically significant; Y = Statistically significant result, where + favours intervention and – favours control

### Experienced shift worker reflections: napping

In nursing, napping for 2 hours prior to the first night shift, and/or napping prior to the end of night shifts can be helpful strategies. Conversely, napping is frowned upon in mining and construction as it is associated with laziness – in fact it can be a sackable offence.

Additionally, although sleep is possible in nursing settings, it is comparatively hard to create an environment where napping could take place in the building sector, even if it was viewed as an acceptable strategy.

### Education and Training interventions (N=3)

Only one of the three studies examining education and training [103] used a referenced measure of mental health (Table 7). Education across the studies encompassed the science of sleep, sleep disorders, and sleep strategies. All studies were pre-post in design with small samples (n=30-61). Although statistically significant findings were reported for a positive effect of education and training on quality of life, burnout and psychological distress, the evidence base is insufficient to draw firm conclusions.

**Table 7:** Summary of studies implementing education and training (n = 3)

| **First Author, Year** | **Study type** | **Population Category** | **Total n participants**<br>Group(s) (description of intervention): n | **Mental health outcomes**<br>**(Sig in bold)** | **Sig. mental health outcome** | **Sig. sleep outcome** |
| --- | --- | --- | --- | --- | --- | --- |
| **Studies using a referenced mental health outcome measure (n = 1)** |  |  |  |  |  |  |
| James et al. 2018 [103] | Pre-Post | Police/Fire/other social services | 61<br>I: fatigue management training | **Quality of Life** | Y+ | Y+ |
| **Studies not using a referenced mental health outcome measure (n = 2)** |  |  |  |  |  |  |
| Holzinger et al. 2019 [104] | Pre-Post | Other shift workers; Trains | 30<br>I: a two-day sleep coaching seminar | **Quality of Life** | Y+ | Y+ |
| Holzinger et al. 2020 [105] | Pre-Post | Other shift workers; Trains | 30<br>I: a two-day sleep coaching seminar | Quality of Life | N | Y+ |

I = intervention group; n = number of participants; N = No statistically significant result (i.e. no difference between groups detected); Statistically significant; Y = Statistically significant result, where + favours intervention and – favours control

### Experienced shift worker reflections: education and training

There is a need to enhance educational interventions in both construction and nursing sectors. In construction, ‘toolbox’ talks can be engaging but still focus on mental health. To reinforce sleep health, in addition to harnessing talks (which are sometimes mandatory prior to commencing building projects), consideration should be given to strategies that support talks, such as posters and opportunities for conversations. Other opportunities to incorporate sleep health education into professional development in construction include ‘rain days’ where work is not possible. In nursing, like construction, there is an existing professional development infrastructure, however awareness of education on sleep is low. It is not incorporated into undergraduate training and although some industry bodies such as unions do produce sleep education it is not routinely incorporated into workplace-based professional development. This identifies an important opportunity for targeted education on sleep health in multiple shift working sectors.

### Other or Multifaceted Interventions (N=11)

Eleven studies examined the effect of combinations of the above interventions or were single studies of an intervention (Table 8). Nine of these used a referenced measure of mental health. Six multifaceted studies examined:

- Education / training + rostering intervention (n=6 healthcare workers, no effect on mood) [115]
- Light + exercise (n=30 aviation workers, positive effect on mood) [106]
- Light + melatonin (n=17 mining workers, no effect on anxiety and depression) [107]
- Light + scheduled sleep (n=39, laboratory-based, no effect on mood) [112]
- Napping + light therapy glasses (n=95, positive effect on wellbeing) [113]
- Zolpidem + napping (n=18 aviation workers, positive effect on mood) [59] Collectively, these studies reported variable effects on mental health outcomes. Given the small sample sizes and variations in intervention combinations, no firm conclusions can be made regarding multifaceted interventions based on these studies.

The remaining five studies examined Transcranial Direct Current Stimulation / Caffeine [109]; Yogic relaxation [110]; Asparagus extract [111]; Homeopathy [114] and Meridian acupuncture [108]. Little confidence can be placed in the findings of individual studies.

**Table 8:** Summary of studies implementing other or multifaceted interventions (n = 11)

| **First Author, Year** | **Study type** | **Population Category** | **Total n participants**<br>Group(s) (description of intervention): n | **Mental health outcomes**<br>(Sig in bold) | **Sig. mental health outcome** | **Sig. sleep outcome** |
| --- | --- | --- | --- | --- | --- | --- |
| **Studies using a referenced mental health outcome measure (n = 9)** |  |  |  |  |  |  |
| Barger et al. 2021 [106] | RCT (crossover) | Aviation workers | **20**<br>I: exposure to blue-enriched polychromatic lighting and exercises before and during shifts | **Mood** | Y+ | N/A |
| Bjorvatn et al. 2007 [107] | RCT (crossover) | Extractive industries;<br>Problems adjusting to shift work | **17**<br>I: 12-h shift workers receiving a placebo, melatonin 3 mg, or bright light exposure (30 minutes) | Anxiety;<br>Depression | N | Y+ |
| Caldwell Jr et al. 1998 [59] | RCT (crossover) | Aviation workers | **18**<br>I: taking 2-h nap (with 10 mg zolpidem or placebo) or 2-h rest without sleep after 38-h wakefulness | **Vigour; tension; depression; confusion; anger; mood** | Y+ | Y+ |
| Cho et al. 2021 [108] | Quasi-experimental; Pre-post | Healthcare workers | **59**<br>C (no intervention): 30<br>I (receiving meridian acupressure on six acupressure points): 29 | **Anxiety; stress** | Y+ | N/A |
| McIntire et al. 2014 [109] | RCT | Military | **30**<br>I1: (tDCS with placebo gum): 10<br>I2: (sham tDCS with caffeine): 10<br>I3 (sham tDCS with placebo): 10 | Mood; Anxiety;<br>Depression; Anger-hostility, Vigor-activity, Fatigue-inertia, Confusion-bewilderment | N | N/A |
| Raghul et al. 2018 [110] | Pre-post | Healthcare workers | **35**<br>I: Performing yoga relaxation (Shavasana) after the day after night shift | **Stress** | Y+ | N/A |
| Sakai et al. 2021 [111] | Pre-post | Healthcare workers; Sleep Problems | **29**<br>I: taken Kenja-no-Kaimin containing asparagus extract prior to sleep for 4 weeks | **Psychological distress** | Y+ | Y+ |
| Smith et al. 2009 [112] | Quasi-experimental | Simulated shift work | **39**<br>I1 (not re-entrainment): 12<br>I2 (partially re-entrained scheduled exposure to bright light and darkness (sleep)): 21<br>I3 (completely re-entrained): 6 | **Mood** | I2: Y+<br>I3: Y+ | Y+ |
| van Woerkom 2021 [113] | Quasi-experimental | Healthcare workers | **95**<br>I: 8-h shift nurses having access to a napping facility, therapy glasses, both facilities or no facilities | **Well-being** | Y+ | Y+ |
| **Studies not using a referenced mental health outcome measure (n = 2)** |  |  |  |  |  |  |
| La Pine et al. 2006 [114] | RCT (crossover) | Healthcare workers | **28**<br>I: night shift nurses randomised to take placebo or a homeopathic tablet | **Quality of Life** | N | N/A |

| First Author, Year | Study type | Population Category | Total n participants<br>Group(s) (description of intervention): n | Mental health outcomes<br>(Sig in bold) | Sig. mental health outcome | Sig. sleep outcome |
| --- | --- | --- | --- | --- | --- | --- |
| Smith-Coggins et al. 1997 [115] | Pre-post | Healthcare workers | **6**<br>Pre: standard shift<br>Post1: standard shift with Ehret and Scanlon diet<br>Post2: forward rotating shift with 2-night shifts maximum and 48-h off duty after a night shift | Mood | N | N |

C = control group; I = intervention group; n = number of participants; N = No statistically significant result (i.e. no difference between groups detected); RCT = randomised controlled trial; Statistically significant; Y = Statistically significant result, where + favours intervention and – favours control

## Discussion

This is the first known systematic review focusing on effectiveness of sleep health interventions on mental health in shift workers. Almost two thirds (n=64) of the 102 included studies evaluated either shift rostering (n=39) or pharmacological interventions (n=25) and over half (n=54) recruited healthcare (n=43) or manufacturing (n=11) workers. Pleasingly, 87 of the 102 included studies used a referenced mental health or mood measure permitting a higher degree of confidence in these studies. The insights from the two experienced shift workers with over 10 years of experience each illustrate the importance of understanding a broad array of workplace contexts given the variable acceptability and feasibility of interventions observed. It is acknowledged that this review did not gather in-depth information across many shift work sectors or examine implementation literature. Hence the insights are not necessarily generalisable to their industries and/or transferable to other sectors. More in-depth co-design with a representative panel of shift workers would be needed to deepen understanding of overarching themes and specific sectoral issues.

### Effective/positive interventions

Rostering studies with large sample sizes demonstrated that a forward rotating shift schedule (i.e., day, afternoon then night shift) has a generally positive impact on mental health compared to the reverse rotation. Several studies in healthcare demonstrated that work hour restrictions in resident-level doctors have positive mental health impacts, and studies in other sectors report shorter shifts are optimal compared with longer shifts (i.e., 10-hours vs. 12+- hours). Hypnotic and stimulant medications have positive mental health effects. Four of the 14 lighting studies also reported positive effects on mood and a further three demonstrated improvements in wellbeing, anxiety and depression. Despite promising findings pertaining to cognitive behavioural therapy, education and training, napping and multifaceted interventions, there were insufficient studies in these categories to warrant firm conclusions.

In many cases, mental health and mood indicators were not the primary focus of the studies. As this review focused on mental health outcomes, in-depth analysis of the full array of outcome measures in the included studies was not conducted. Future analysis of the balance between mental health and sleep outcomes, as well as exploration of the links between these at the level of individual studies, would shed light on this important interaction.

The perspectives of two experienced shift workers enhanced understanding of areas in which there is little or no research, yet which are explored in real-world settings. Interventions used by these members of the research team (MA and DF) yet not explicitly represented in the 102 studies include alterations in dietary habits (including reducing alcohol consumption), use of air conditioning in hot climates, use of wearable technologies and ear plugs and white noise to distract from daytime interruptions to sleep. Conversely, the experienced shift worker researchers were less familiar with some interventions such as the use of goggles or sunglasses to avoid light at inappropriate times.

Other insights and nuances from experienced shift workers also sharpen the focus on review findings. For example, there are clear opportunities to better promote the importance of sleep hygiene among shift workers. The need to better promote sleep health in nursing, for instance, was surprising given the amount of research that has been conducted in this sector. This implies that there is still much work to be done in translating research knowledge into policy and practice. Although some studies have examined the consequences of shift work on social activity, the role of family in supporting the demands of shift workers is under-researched.

Another important but under-represented consideration pertains to commutes home from shift work. For example, on selected building projects, any employee with a commute duration of an hour or more is offered nearby accommodation for recovery prior to the commute. This is consistent with research indicating that driving for 45 minutes or more following a night shift may be unsafe [116].

There are important cultural differences between work sectors in relation to sleep health interventions. Where mining and construction workers have relatively high buy-in to structured education such as ‘toolbox’ talks, updates of in-service education can be low in nursing, with older nurses in particular benefitting more from informal ‘hallway’ conversations; napping is facilitated in nursing but frowned upon in construction; lighting is less practically applicable in construction compared to nursing. This underscores the importance of sector-specific approaches to research and practice.

Shift work by its nature restricts access to healthcare services such as general practitioners. This means that they are less likely to be exposed to this cohort, and potentially less aware of circadian rhythm disturbances and the specific interventions that may address them. Limited access to healthcare services also reduces opportunities for shift work employees to identify and manage mental (and general) health issues as well as sleep disturbances. A possible strategy for addressing this is organisational rather than community-level provision of health support services, either in-person (for example onsite nurses and other professionals), or via other resources including online platforms.

### For who

A major limitation in the literature is a lack of studies examining mental health outcomes in occupational groupings outside of healthcare, manufacturing, emergency services and mining. These include transport and logistics, call centre workers and cleaners. Even amongst the occupations studied, the health workforce was dominant. This is the only occupational cohort in this review where firm conclusions can be drawn regarding the effect of sleep interventions on mental health, in particular with regards to rostering and pharmacological interventions which were the most frequently studied in the review.

### In what context

The included studies were predominantly undertaken in first-world settings, notably the United States and Europe. Studies of shift workers in Low and Middle-Income Countries (LMIC) were lacking. Workplace regulatory standards for workers in LMIC countries are potentially less rigorous; for example, the two biggest influences on work hour restrictions in doctors were the US Graduate Medical Education resident duty hour limits and the European Working Time Directive on junior doctors. Knowledge of similar regulations in LMIC is lacking, and no research examining duty hours was identified in this review.

Strengths of this review are a comprehensive search strategy across multiple databases; independent screening of all citations / abstracts and full text studies by two researchers; and a broad definition of interventions to ensure that all primary studies measuring mental health outcomes were captured. There are some limitations to acknowledge in this version of the review. Although there were no year restrictions on the search, non-English studies were excluded. Whilst study design, sample size and use of a referenced mental health outcome measure were used to interpret the findings, no formal quality appraisal was undertaken owing to the large volume and heterogeneity of the studies.

## Recommendations

The most critical recommendation for business leaders, policymakers and researchers is that mental health impacts and outcomes associated with shift work require greater recognition. Sleep health interventions for shift workers appropriately focus on influencing circadian timing, sleep length and sleep quality. Improvements in these parameters are also likely to positively influence mental health because of the known link between poor sleep, circadian rhythms, and mental illness.

Relatedly, more consideration needs to be given to multifaceted interventions that combine strategies addressing both sleep and mental health. This is especially important given the large array of strategies reported by the experienced shift workers in the research team.

Research into the effectiveness of such approaches is lacking; only six of the 102 included studies examined multifaceted interventions.

There is presently little or no research addressing the majority of shift working populations around the world. More research examining mental health outcomes must be directed at occupational groups outside of health and in LMIC settings. Although the focus on healthcare shift workers has appropriately been driven by knowledge of burnout in the medical professions as well as the risk to patients, equal or even greater risks exist, for example in mining and transport and in poorly regulated occupational settings.

Finally, despite a large number of studies being identified, most studies had small sample sizes. This reduces confidence in and transferability of findings, even when robust study designs are employed. Multi-centre studies spanning larger populations and even countries are therefore needed. This is feasible given the reproducibility of interventions such as setting shift schedules, using pharmaceuticals or applying light therapy. Better co-ordination and standardisation of such interventions would also address the heterogeneity within each intervention category, which further compromises applicability of identified studies.

## Conclusions

A systematic review of sleep health interventions for managing mental health in shift workers identified 102 primary studies. Interventions predominantly examined shift rostering (n=39), pharmacological therapies (n=25) and used of light (n=14). Healthcare workers were the focus of almost half (n=43) of the included studies. Utilising forward-rotating shifts (day, afternoon, then night shift) and restricting work hours for junior doctors were shown to positively impact mental health and mood outcomes; however, a small number of studies also reported negative impacts of hour restrictions. Both sedative and stimulant medications were also shown to have positive mental health effects. There was evidence supporting the use of lighting interventions to enhance mental health, however sample sizes were small. Few conclusions could be drawn on other interventions. Based on the perspectives of two experienced shift workers, there is considerable variation in the acceptability and feasibility of interventions across different sectors. This is driven in part by sector-specific cultural norms and relatively low awareness of sleep hygiene and its relationship to mental health.

Given these insights, the dearth of research addressing mental health outcomes outside of the health professions assumes even greater importance. There is also a lack of research beyond high-income countries. Many studies did not give the same weight to mental health and sleep outcomes, as reflected by lack of multifaceted intervention studies. With the majority of the global shift working population under-represented by existing research, this area warrants major focus by business leaders, policy makers and researchers.

## Data Availability

All data produced in the present study are available upon reasonable request to the authors.

## Acknowledgements

The author team thank Katrina Tepper from the Monash University Library for her expert assistance in developing the search strategy for this review; and Cong Lem for his assistance in data extraction.

This project was funded by The Wellcome Trust.

The authors have no conflicts to declare.

