## Supplementary File 1 for "Sleep health interventions for managing mental health in shift workers: A systematic review"

**Supplementary File 1: Search Strategies**

**String 1: Population - shift workers**

1. ((shift OR shifts) adj1 (rota OR system OR systems OR schedul* OR hours OR time OR pattern$ OR cycle OR extend$ OR evening OR late OR roster OR early OR weekend OR twilight OR graveyard OR night$ OR split OR non-standard OR "non standard" OR flex$ OR turnaround OR continuous OR rotat$)).tw.

2. (day adj2 schedule?).tw.

3. (rota OR roster OR 'day week' OR flexitime OR 'hours of work' OR nightshift* OR shiftwork*).tw.

4. ((work$ OR duty) adj1 (shift OR shifts OR rota OR system OR systems OR schedul* OR hours OR time OR pattern$ OR cycle OR extend$ OR evening OR late OR roster OR early OR weekend OR twilight OR graveyard OR night* OR split OR non-standard OR "non standard" OR flex$ OR turnaround OR continuous OR rotation$)).tw.

5. ((backward OR forward OR rapid OR slow OR rapidly OR slowly OR advancing OR delaying) adj1 (rotation OR rotate OR rotating)).tw.

6. (rota OR roster OR duty OR shift OR shifts OR shiftwork OR hours OR week OR work).mp.

7. 5 and 6

8. 1 OR 2 OR 3 OR 4 OR 7

Where available: "Shift Work Schedule"[MeSH]

**String 2: Condition**

Sleep OR “Sleep disturbance*” OR “sleep wake disturbance*” OR “sleep disorder*” OR sleepiness OR "sleep initiation and maintenance disorders" OR circadian OR “circadian rhythm disturbance” OR “circadian rhythm” OR “circadian rhythm misalignment” OR fatigue OR “poor-quality sleep” OR “inadequate sleep” OR “impaired alertness” OR “alertness impairment*” OR “Chronobiology Disorder*” OR vigilance OR alertness OR alert OR wakefulness OR drowsiness OR fatigue OR insomnia OR hypersomnolence OR dyssomnia OR eveningness OR morningness OR "concentration difficulties" OR attentiveness OR arousal OR performance OR vigilance OR vigilant OR “sleep quality” OR chronotype

**String 3: Intervention**

Intervention OR trial OR pilot OR study OR program* OR strateg* OR activit* OR incentiv* OR manage* OR control OR address OR experiment* OR campaign* OR initiative* OR OR promote* OR promotion OR foster* OR motivat* OR engage* OR behav* OR “behavio$r change”

**String 4: Outcomes**

stress OR burnout OR anxiety OR depression OR “mental health” OR “mental disorder*” OR “mental illness*” OR psychological OR “quality of life” OR wellbeing OR well-being OR “quality of working life” OR “personal satisfaction” OR “work satisfaction” OR “life satisfaction” OR “work functioning” OR “work* standard*” OR “happiness” OR mood

**1 AND 2 AND 3 AND 4**
