## Supplemental File 2 for "Sleep health interventions for managing mental health in shift workers: A systematic review"

**Supplementary File 2: Invitation for recruitment of lived experience researcher**

**Sleep health interventions for preventing and managing mental illness in the workplace**

**Invitation to join research team as a lived experience team member**

To whom it may concern,

We are seeking shift workers with lived experience of mental illness associated with sleep disturbance to join a small research team applying for funding to undertake a research literature review in this area.

[The Wellcome Trust based in the UK](https://wellcome.org/what-we-do/our-work/mental-health-transforming-research-and-treatments#commissions-dce9) are funding 20 research teams from around the world to provide an insight analysis into a promising approach for preventing or addressing mental health problems in the workplace.

Our research team submitted an initial expression of interest to this funding opportunity with a proposal focus on managing sleep in shift workers. From an initial pool of 120 applicants, we have been shortlisted as one of 40 teams invited to submit a full application for funding. If successful, our research review project will run from **October 18, 2021, to March 21 2022**

**Details of the role:**

As a member of the research team, we would require you to participate in the following activities:

- Attend research team meetings to develop the research protocol, discuss progress and allocate tasks (approximately 2 hours per month = 8 – 10 hours)
- Provide reflections on the feasibility, acceptability and potential impact of interventions in the research literature, based on your lived experience (approximately 10 hours from Dec 2021 – Feb 2022)
- Contribute to the writing of reports and presentations arising from the review, especially to aid interpretation of what the review findings mean for practice based on your lived experience (approximately 5 hours from Jan 2022 – March 2022)

**Remuneration:**

This is a paid project role. We will remunerate you at your hourly rate in your current or last primary paid employment role + 20%.

Thank you for considering this role. To discuss your involvement further please or call me on **0403 197 275**.

Yours Sincerely,

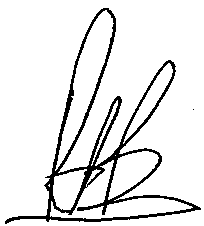

Associate Professor Peter Bragge, Lead Investigator

on behalf of the research team:

- Professor Shantha Rajaratnam
- Professor Jane Burns
- Dr Tracey Sletten
- Ms Alyse Lennox
