## Supplementary File 3 for "Sleep health interventions for managing mental health in shift workers: A systematic review"

**Supplementary file 3: Results of study selection: Prisma flow diagram**

**7,006** studies identified through database searching

**2,106** duplicates removed

**102 primary studies included**

**341** studies excluded

- **95:** Review (72 published prior to 2017; 23 published after 2017 retained for reference list scanning)
- **55**: Did not measure mental health outcomes
- **49**: Conference abstract
- **38**: Ongoing studies (16 published prior to 2017 including one pilot; 22 published from 2017 retained for cross-checking against included studies)
- **28**: Observational not interventional study design
- **17**: Duplicate
- **17**: Non-English
- **13**: Study population is not majority shift-workers
- **11**: Not a sleep health intervention
- **5**: No full text available
- **8**: Commentary
- **2**: Media article
- **2**: Thesis / dissertation
- **1**: Animal model

**443** full-text studies assessed for eligibility

**4,457** studies excluded based on title & abstract

**4,900** studies screened
